# Community Mobility and COVID-19 Dynamics in Jakarta, Indonesia

**DOI:** 10.1101/2021.07.24.21261016

**Authors:** Ratih Oktri Nanda, Aldilas Achmad Nursetyo, Aditya Lia Ramadona, Muhammad Ali Imron, Anis Fuad, Althaf Setyawan, Riris Andono Ahmad

**Author notes:** These authors contributed equally to this work (Ratih Oktri Nanda, Aldilas Achmad Nursetyo, Aditya Lia Ramadona, Althaf Setyawan). These authors also contributed equally to this work (Muhammad Ali Imron, Anis Fuad, Riris Andono Ahmad).

## Abstract

**Background:** Human mobility could act as a vector to facilitate the spread of infectious diseases. In response to the COVID-19 pandemic, Google Community Mobility Reports (CMR) provide the necessary data to explore community mobility further. Therefore, we aimed to examine the relationship between community mobility on COVID-19 dynamics in Jakarta, Indonesia.

**Methods:** We utilized the mobility data from Google from February 15 to December 31, 2020. We explored several statistical models to estimate the COVID-19 dynamics in Jakarta. Model 1 was a Poisson Regression Generalized Linear Model (GLM), Model 2 was a Negative Binomial Regression Generalized Linear Model (GLM), and Model 3 was a Multiple Linear Regression (MLR).

**Results:** We found that Multiple Linear Regression (MLR) with some adjustments using Principal Component Analysis (PCA) was the best fit model. It explained 52% of COVID-19 cases in Jakarta (R-Square: 0.52, p<0.05). All mobility variables were significant predictors of COVID-19 cases (p<0.05). More precisely, about 1% change in grocery and pharmacy would contribute to a 4.12% increase of the COVID-19 cases in Jakarta. Retails and recreations, workplaces, transit stations, and parks would result in 3.11%, 2.56%, 2.26%, and 1.93% of more COVID-19 cases, respectively.

**Conclusion:** Our study indicates that increased mobility contributes to increased COVID-19 cases. This finding will be beneficial to assist policymakers to have better outbreak management strategies, to anticipate increased COVID-19 cases in the future at certain public places and during seasonal events such as annual religious holidays or other long holidays in particular.

## Introduction

The novel coronavirus, which was further called SARS-CoV-2, first emerged in November 2019 in Wuhan, China. The epidemic subsequently began in December 2019, and the cases were rising in January 2020. On January 30, the World Health Organization (WHO) declared the pandemic “Public Health Emergency of International Concern.” Around eight months after cases were found in China, the pandemic has not yet been contained and impacted more than 200 countries across continents in total[1].

Human mobility act as a vector for the spread of infectious agents. Therefore, mobility restriction is a common strategy to slow down the pandemic. Some countries such as China and New Zealand have already demonstrated the success of the epidemic containment by putting strict social distancing measures as the priority in the early stage of the epidemic[2, 3]. However, different policies in some countries and some other factors contributed to the longer duration of the pandemic in several countries, such as Indonesia.

Previous studies of H1N1 reported that population mobility could be a good prediction for the epidemic spread in international travel [4]. In response to the COVID-19 pandemic, various mobile-phone data on population movement were publicly made available. The previous study conducted in the early stage of the pandemic has shown the potential of assessing mobility data in building epidemiological models to anticipate COVID-19 spread.[5] Moreover, it is reported that COVID-19 spread was explained by the population mobility gathered from various mobility data sources, that the reductions in mobility would contribute to decreased COVID-19 cases. [6, 7]

Google Community Mobility Reports (CMR) is a mobility data source that features population movement patterns in several categories; retails and recreations, groceries and pharmacy, transits stations, parks, workplaces, and residential. Several studies reported utilizing CMR for cross-country comparisons[8], national levels[9] and sub-national levels[10]. Earlier study also reported that the strategy which emphasizes the decrease of public places mobility is effective to slow down the pandemic [7].

In Indonesia, the government did not enforce a national lockdown policy. Instead, they implemented partial mobility restrictions. The partial mobility restriction, known as large-scale social restrictions (PSBB), was implemented depending on local epidemiological status. In Jakarta, the capital city and the epicenter of the early COVID-19 epidemic in Indonesia, different mobility restrictions were implemented from early March until November 2020. As one of the most populous cities in Indonesia, the provincial government of Jakarta implemented PSBB to limit people’s movements by postponing public events, limiting transportation (MRT, LRT, and bus) routes and their capacity. The government also closed schools and other public places that might induce large gatherings and put into effect the work-from-home policy in specific fields [11]. However, COVID-19 cases in Jakarta were still rising to the end of the year, exceeding 180,000 by December 30 [12]. PSBB and its relaxation (PSBB Transisi), along with several events (long-weekend, religious holidays), might contribute to increased mobility within the city. Less is known about how the movement of the people within the city influenced COVID-19 cases in Jakarta and its further implications for local regulation.

This study aimed to explore how human mobility observed from CMR impacts COVID-19 dynamics in Jakarta. The reported mobility data acts as a reflection of the implementation of mobility restrictions and other occurring events. We hypothesize that the increase in population mobility is related to increased COVID-19 cases.

## Materials and Methods

### Data on cases of COVID-19

We conducted a cross-sectional study. We used COVID-19 data in Jakarta provided by the Kawal COVID-19 website due to the discrepancy between the regional health office and the health ministry. Kawal COVID-19 is an independent website that provides information and collects data on Indonesia’s COVID-19 pandemic from the government at the provincial levels. Thus, the data is more reliable to use [12]. We use the data from February 15 to December 31, 2020.

### Data on community mobility

We retrieved the mobility data from the Google Community Mobility Report (CMR) Data. Google collected anonymized users’ location history data [13]. The data shows the changes in visits to places categorized as retails and recreations, groceries and pharmacies, parks, transit stations, workplaces, and residential. The data is collected by considering the number of requests made to Google maps for directions in several countries, sub-regions, cities, and minimum thresholds for direction requests per day. The report then shows the percentage of mobility change in places such as retails and recreation, groceries and pharmacy, parks, workplaces, transit stations, and residential. The descriptions of the places are as follows:

- Retails & Recreational: mobility trends for places like restaurants, cafes, shopping centers, museums, libraries, and picture theatres
- Grocery & Pharmacy: mobility trends for places such as grocery, food warehouses, markets, local hats, farmer’s markets, specialty food shops, different drug or medicine stores, and pharmacies
- Parks: places of attraction like local parks, national parks, public beaches, marinas, dog parks, plazas, and public gardens
- Transit stations: mobility trends for places like public transport hubs such as subway, bus, and train stations
- Workplaces: mobility trends for places of work
- Residential mobility: mobility trends for places of residence

We applied statistical transformations to our data and used STATA software 14.0 to analyze the data. We used the five variables in Google Community Mobility Reports (retails and recreations, groceries and pharmacies, parks, transit stations, and workplaces) as the study exposures and the observed COVID-19 new cases as the study outcome. We excluded residentials as we assumed it explained immobility rather than mobility.

To determine lag, we used a cross-correlation function in R. Moreover, to reduce the weekend-weekday bias of COVID-19 cases data counting, we applied 7-Days Moving Average (7DMA). Regression models were used to predict the continuous dependent variable. In this paper, the dependent variable is COVID-19 cases. We first calculated the 7-Day Moving average of the dependent variable. After analyzing the cases data, we found that the data had a skewed distribution. Therefore, we did the log-transformation to the data to follow the normal distribution to fit our model better. For the independent variables, we have our data lagged by seven days.

We explored several models to estimate COVID-19 cases. We included Poisson Regression GLM and Negative Binomial GLM in the analysis as both analyses were suitable for modeling count data. Additionally, we also added Multiple Linear Regression (MLR) for the comparison. Therefore, we compared a Poisson Regression GLM (Model 1) with a Negative Binomial Regression GLM (Model 2) and an MLR (Model 3). We used three parameters of R-Square, Akaike Information Criterion (AIC), and Root Mean Square Error (RMSE) to assess the best fit model. A larger value of R-Square indicates the better the model fits the observations. Meanwhile, it is the lower values of AIC and RMSE that indicate a better-fit model.

### Variable selection

The mobility types recorded in the Google CMR are not independent of each other, e.g., people might go to workplaces, groceries, and public transit in a day, multicollinearity of the data exists. However, having different combinations of mobility gives different levels of exposure. Therefore, we applied Principal Component Analysis (PCA) to consider the contribution of different types of mobility to the overall mobility exposures.

## Results

To determine lag days applied to the dataset, we use a cross-correlation function. This function applied Pearson Correlation to the independent variables while applying lag at the same time. The result of cross-correlation function analysis can be seen in **Figure 2**. Note that four variables have a higher correlation on negative lag. Therefore, combined with assumptions that COVID-19 will develop symptoms within 7-14 days, we take seven days as lag applied to the variables.

We initially checked the assumptions of the linear regression and found some multicollinearity between the independent variables by the given p-value. Based on the R-Square values, we found that the MLR model, where all of the mobility variables included, explained the variance by 66.39%. However, the initial result of the Multiple Linear Regression showed that groceries and pharmacies (P < 0.05), transits stations (P < 0.05), along with workplaces (P < 0.05) are significant. Thus, multicollinearity existed. **(Table 1)**

**Table 1.**
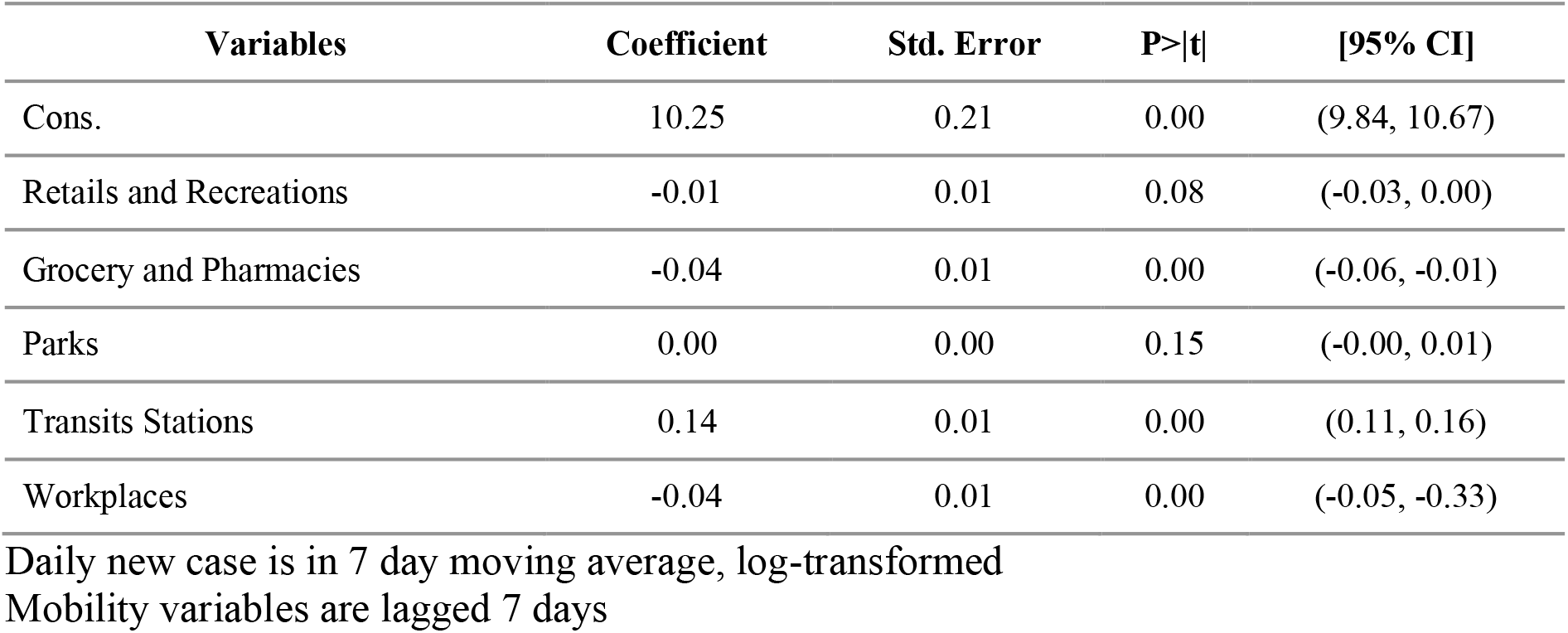
Multiple linear regression analysis of the mobility variables and COVID-19 daily new case

| Variables | Coefficient | Std. Error | P>\|t\| | [95% CI] |
| --- | --- | --- | --- | --- |
| Cons. | 10.25 | 0.21 | 0.00 | (9.84, 10.67) |
| Retails and Recreations | -0.01 | 0.01 | 0.08 | (-0.03, 0.00) |
| Grocery and Pharmacies | -0.04 | 0.01 | 0.00 | (-0.06, -0.01) |
| Parks | 0.00 | 0.00 | 0.15 | (-0.00, 0.01) |
| Transits Stations | 0.14 | 0.01 | 0.00 | (0.11, 0.16) |
| Workplaces | -0.04 | 0.01 | 0.00 | (-0.05, -0.33) |

Daily new case is in 7 day moving average, log-transformed Mobility variables are lagged 7 days

To address this issue, we used the Principal Component Factor Analysis to develop a new composite variable. The three variables that showed multicollinearity (grocery and pharmacies, transits stations, and workplaces) were combined into a composite index using a z-score. To this end, we created five models which consisted of different predictors. The first four models consisted of parks and retail, with the addition of grocery and pharmacy, transits stations, and workplaces in the model separately. The last model was built from a composite of grocery & pharmacy, transit stations, and workplaces, then combined with the rest of the independent variables (parks and retails and recreations).

We explored the model, which consisted of different predictors. As seen in **Table 2**, Multiple Linear Regression showed a better result in terms of lower AIC and RMSE and higher R-Square compared to Poisson Regression and Negative Binomial Regression.

**Table 2.** Poisson GLM, Negative Binomial GLM, and Multiple Linear Regression of COVID-19 Daily New Cases

| Model | AIC |  |  | R-Square |  |  | RMSE |  |  |
| --- | --- | --- | --- | --- | --- | --- | --- | --- | --- |
|  | Pois | NB | MLR | Pois | NB | MLR | Pois | NB | MLR |
| 1. Parks_Retails | 3.75 | 5.73 | 2.33 | 0.47 | 0.49 | 0.49 | 0.78 | 0.78 | 0.77 |
| 2. Parks_Retails_Grocery | 3.76 | 5.74 | 2.34 | 0.50 | 0.49 | 0.50 | 0.78 | 0.78 | 0.77 |
| 3.Parks_Retails_Transits | 3.74 | 5.74 | 2.15 | 0.58 | 0.58 | 0.59 | 0.71 | 0.71 | 0.70 |
| 4.Parks_Retails_Workplaces | 3.76 | 5.74 | 2.31 | 0.50 | 0.50 | 0.51 | 0.77 | 0.77 | 0.76 |
| 5. Parks_Retails (Z-Score Grocery_Transits_Workplaces) | 3.76 | 5.74 | 2.29 | 0.51 | 0.51 | 0.52 | 0.76 | 0.76 | 0.76 |

All independent variables are lagged 7 days, composite variables: grocery and pharmacies, transits stations, and workplaces. The dependent variable is in 7-day moving average and log-transformed.

Furthermore, Model 3 with parks, retails and transits had lower AIC and RMSE and higher R-Square value. Model 5 consisted of a composite index of three significant variables (grocery and pharmacy, transit stations, and workplaces), putting it into an analysis along with parks and retails and recreation. This model then explained the variance in COVID-19 cases by 52%, with a lower AIC value (2.29), and RMSE value (0.76). It is the most suitable model that could provide a better explanation. Hence, we considered choosing this model to be analyzed further in this study.

Based on the result of the Multiple Linear Regression analysis in **Table 3**, the equation is as follows:

**Table 3.** Multiple linear regression analysis using daily new case and mobility variables

| Variables | Coef. | Std. Err. | P > \|t\| | 95% CI |
| --- | --- | --- | --- | --- |
| Cons. | 8.44 | 0.28 | 0.00 | (7.88, 9.00) |
| Z_Score_Grocery_Transits_Workplaces (Lagged 7 days) | 0.41 | 0.12 | 0.00 | (0.18, 0.65) |
| Parks (Lagged 7 days) | 0.02 | 0.00 | 0.00 | (0.01, 0.02) |
| Retails and Recreations (Lagged 7 days) | 0.03 | 0.00 | 0.00 | (0.01, 0.45) |

The dependent variable is in 7-day moving average, log-transformed

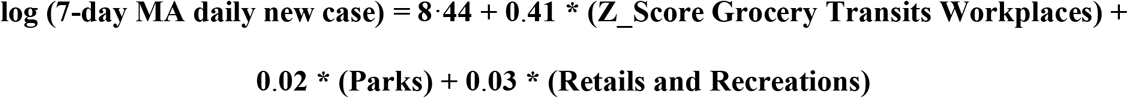

Due to the log transformation of the cases as the dependent variable, and using a composite index (z-score) of three mobility variables, further steps were needed to interpret the results in terms of the effect of each mobility variable to the cases. The composite index of three mobility variables need to be inserted into a division with standard deviation of each of them as the denominator, and the result to be exponentiated **(Table 4)**. The same equation needed to be done to the confidence interval. The coefficient of the other two mobility variables, parks, and retails and recreation, only need to be exponentiated.

**Table 4.** Regression coefficient of mobility variables.

| Variable | Coef. (exp) | Coef. (Percentage) | Mean | Std. Dev | 95% Confidence Interval (Exp) |
| --- | --- | --- | --- | --- | --- |
| Grocery and pharmacies | 1.0412 | 4.12 | -12.30 | 10.34 | (1.01, 1.06) |
| Transits stations | 1.0226 | 2.26 | -42.98 | 18.64 | (1.00, 1.03) |
| Workplaces | 1.0256 | 2.56 | 30.01 | 16.54 | (1.01, 1.04) |
| Parks | 1.0193 | 1.93 | -57.22 | 24.02 | (1.01, 1.02) |
| Retails and recreation | 1.0311 | 3.11 | -34.03 | 16.03 | (1.01, 1.04) |

To enhance the interpretation, we subtracted each of the mobility variables with one, to be subsequently multiplied by a hundred percent. Accordingly, we found that Grocery and Pharmacies is the highest among all mobility variables, whereas one percent increase in Grocery and Pharmacies mobility contributed to 4.12% increase of the cases. Retails and Recreations mobility contributed to 3.11% increase. Transits stations and workplaces contributed to the increase of cases by 2.26% and 2.56% respectively. Meanwhile, parks accounted for 1.93% rise in the cases **(Table 4)**.

### COVID-19 Cases and Containment in Jakarta

The mobility changes as seen in **Figure 1** showed that the changes in population mobility trend are in line with the existing restriction policy and regulations and the holidays. The government revised Jakarta’s mobility restrictions several times to adjust to COVID-19 dynamics from February 15 until December 31 2020. This includes the large-scale social restrictions (PSBB), followed by the relaxation (PSBB Transisi), shifting back to the tighter restrictions (PSBB), and back in relaxation (PSBB Transisi and PSBB Transisi Extended) as the cases seemed to be dropping down. **(Figure 3)** Several events, such as religious holidays (Eid Al-Fitr), public holidays, where some lead to long weekends, are observed, causing the community mobility changes and increased COVID-19 cases afterwards.

**Fig 1.**
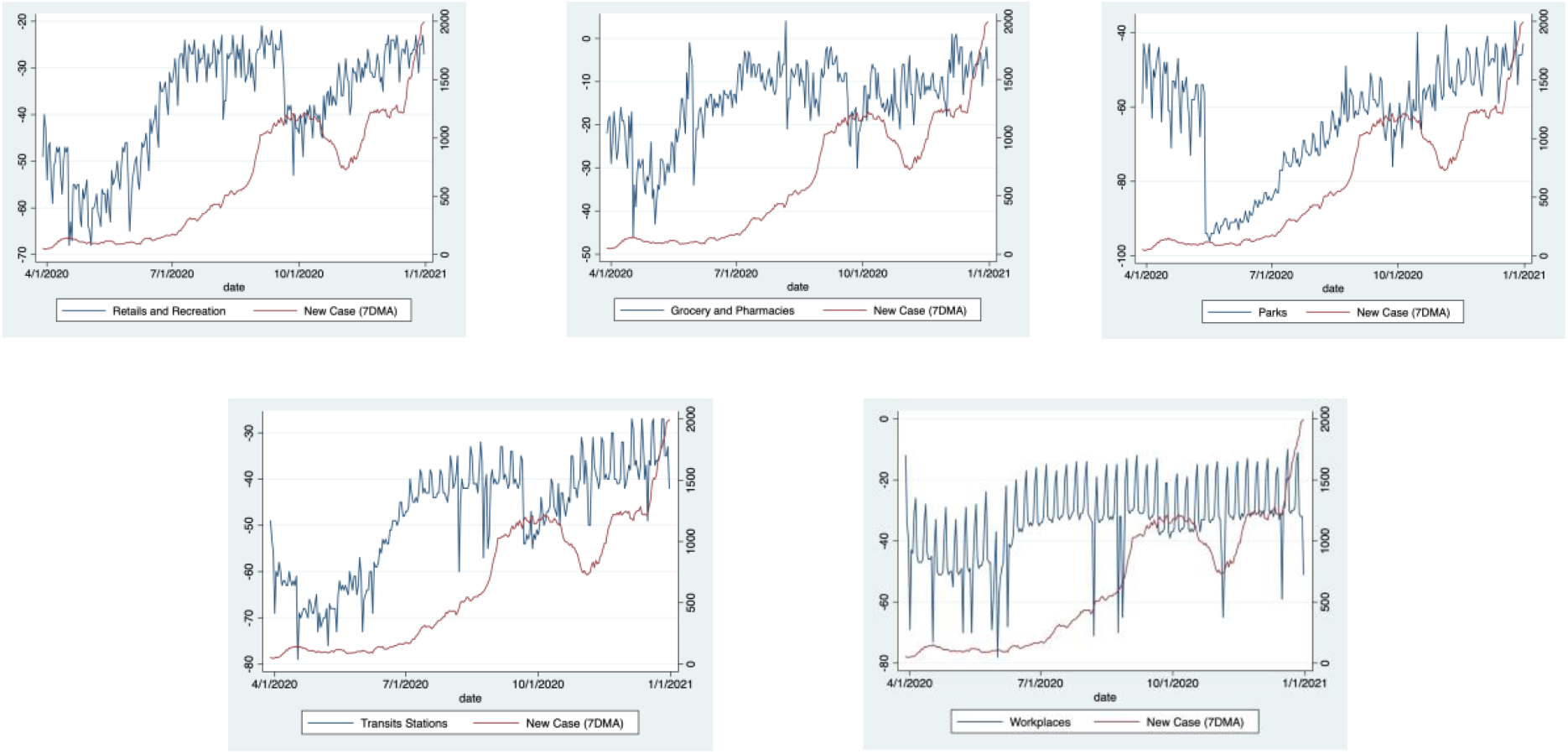
Changes in Human Mobility in Jakarta, 2020.

**Fig 2.**
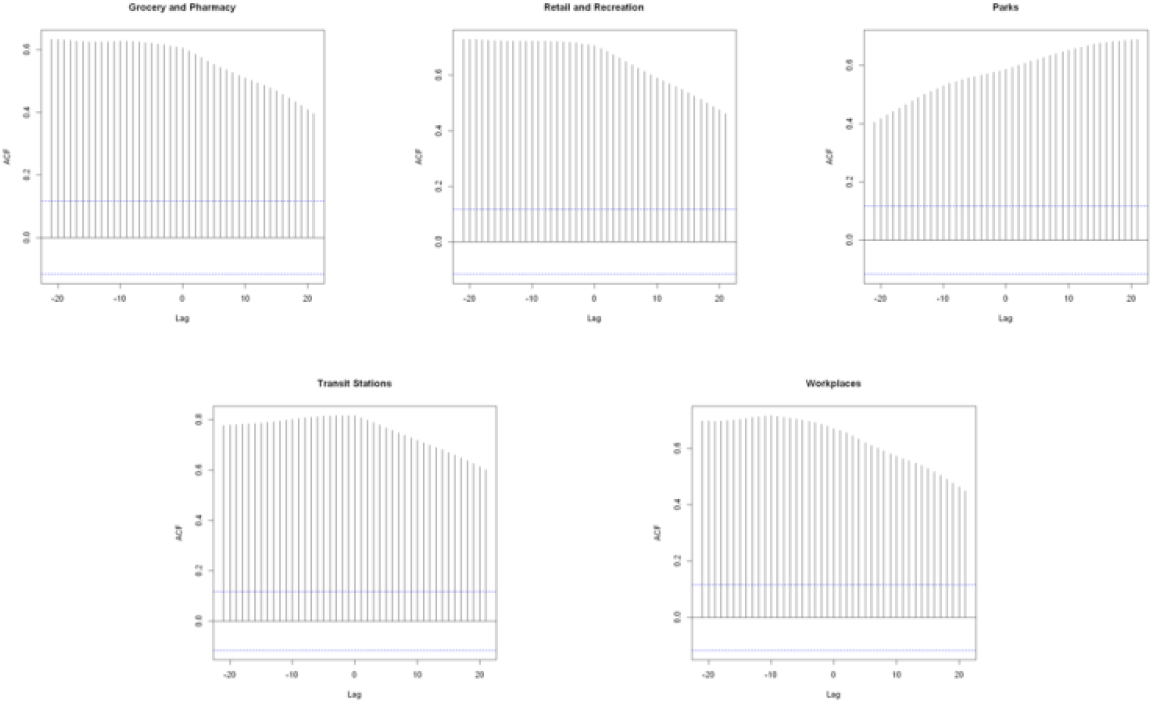
Correlation plot between independent variables and daily confirmed case variables at different time lag.

**Fig 3.**
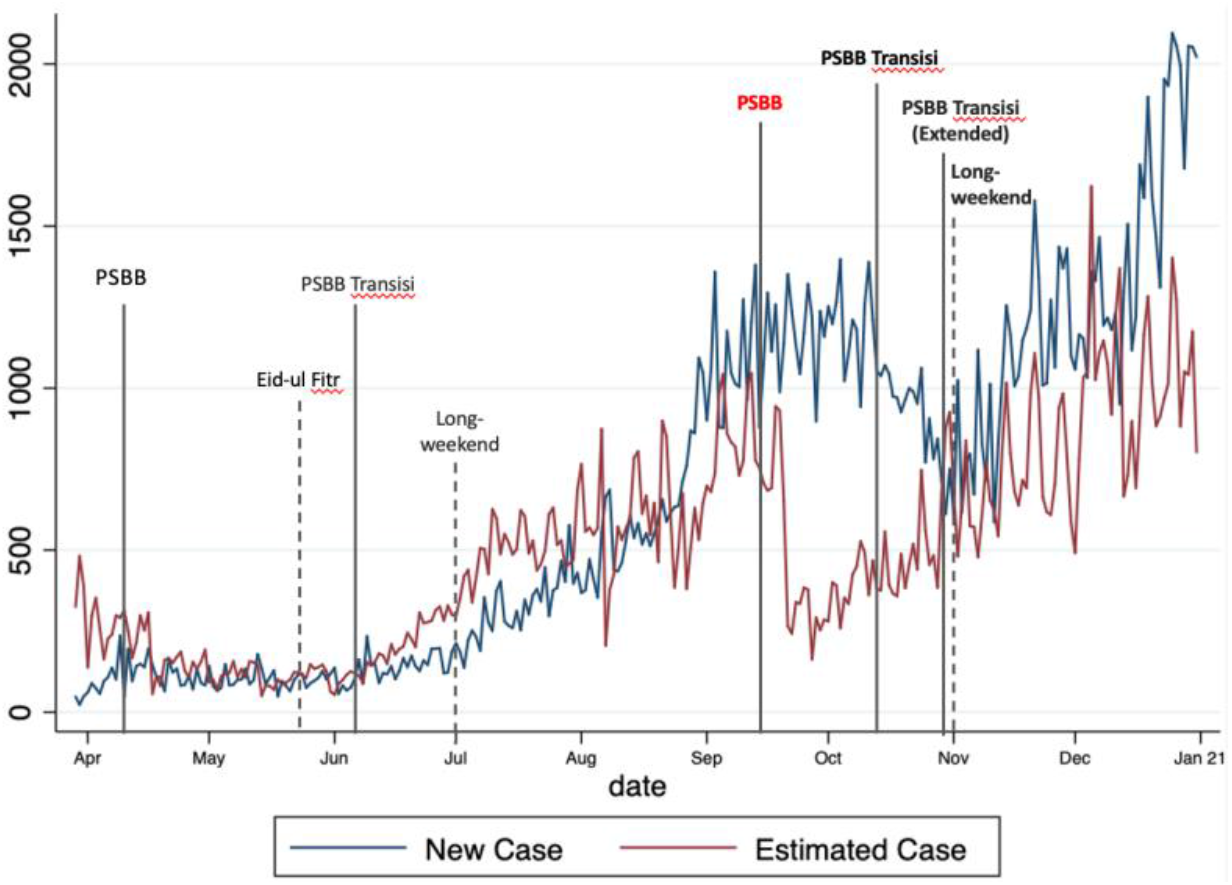
Observed and estimated values of COVID-19 cases.

In Figure **3**, where the observed (actual cases) and predicted values of COVID-19 new cases based on the model was shown, a gap in the period of middle-to-late September towards late November appeared, when PSBB. We assumed that this gap occurred due to the nature of the data, where a decrease of changes in community mobility during the same period was found, where almost all of the mobility category showed a decline in trends **(Figure 1)**. This model, however, does not use time-dependent data. Therefore, when the mobility decline is found in the period starting from the middle of September (PSBB), the model estimated the decline in cases.

## Discussion

### Effect of mobility by categories on COVID-19 dynamics

This study explored the influence of population mobility on COVID-19 dynamics in Jakarta, Indonesia. We demonstrated that the different types of community mobility (retails and recreation, groceries and pharmacies, parks, transit stations, and workplaces) partially explain theCOVID-19 dynamic in Jakarta. Furthermore, we found that mobility in groceries and pharmacies, followed by retails and recreation, transit stations, workplaces, and parks, contributed to the COVID-19 dynamics.

The model we used in this study demonstrated the relationship between mobility and COVID-19 dynamics with the fewest assumptions possible, considering mobility categories as the explanatory variable. We used Multi Linear Regression to explore the impact of community mobility on COVID-19 dynamics in Jakarta. This model has anticipated underreporting or overreporting COVID-19 cases reports by transforming the daily cases into moving average. In terms of mobility data, As the mobility types recorded in the Google CMR are not independent of each other, e.g., people might go to workplaces, groceries, and public transit in a day, multicollinearity of the data exists. However, having different combinations of mobility gives different levels of exposure. Therefore, we applied Principal Component Analysis (PCA) to consider the contribution of different types of mobility to the overall mobility exposures. Additionally, some confounding and other possible limitations might exist during the data exploration. Therefore, the result in this study should be interpreted with caution.

Mobility on groceries and pharmacies contributed the highest in the COVID-19 new cases increase. Food warehouses, farmers’ markets, specialty markets, and drugstores were among the essential businesses allowed to be opened during the restrictions. This finding is in line with a previous study in India, where travel for daily needs purposes are related to COVID-19 transmission.[14] A previous study observed that people shifted from restaurants towards groceries and food sellers during the stay-at-home orders [15]. We assumed that markets, mainly traditional markets, were places with frequent visitors in Jakarta as those places supplied daily necessities for the people. However, traditional markets tend to be crowded and unorganized, also challenging for social distancing implementation. Though the government ruled out health protocols in traditional markets, it was not strongly enforced. Around 107 traditional market clusters and 555 cases were reported in Jakarta, only in the first relaxation period. 22 Precisely, traditional markets contributed 4.3% of increased COVID-19 new cases in Jakarta [16].

Places like café, restaurants, shopping malls, recreation areas, and parks in Jakarta were shut down in March-June and subsequently reopened, adjusting around 50% of the total capacity during the first relaxation period, inducing high mobility to those places. Earlier studies have found that virus transmission most likely occurred in indoor environments, most precisely in crowded and poorly ventilated areas [17]. Hence, without interpersonal distance and ventilation improvement, there is a strong possibility of the airborne transmission of COVID-19 in the retails and recreations sector [18].

At the beginning of the pandemic, the government of Jakarta ruled out the tight restrictions by limiting the number of passengers in public transportations by implementing the SIKM (Exit-Entry Access) from and to Jakarta to reduce the transmission within the city and to other cities or provinces. However, as the restrictions were relaxed, followed by the reopening of the public places, and in the events of religious and public holidays, the visits to transits stations within the city increased. As a result, transit stations (bus, MRT, LRT, etc.) generally possess a risk of crowding. Meanwhile, confined and closed environments in public transportations have a high risk of infectious disease transmission [19]. In terms of workplaces, the distinction of mobility was not very clearly observed. Some people were still working in the informal sectors, and some office workers were still required to work from the office, thus explaining the minor fluctuations trends in this sector.

### Mobility relaxation, seasonal events, and COVID-19 dynamics

Throughout the year, there are many public holidays, including religious holidays, and some of those coincidentally lead on the long weekends. One of the biggest religious events, Eid-ul Fitr, has been predicted to induce the spike of population mobility due to the migrations of people known as homecoming (mudik). The government did enforce the travel restrictions to anticipate a significant spike of COVID-19 cases due to high travels, yet COVID-19 cases increased a few weeks after the event.

Complete closure and limitation of public places visited at the beginning of the pandemic would limit population movement and reduce COVID-19 cases. A study found that the first mobility restrictions regulations implemented in Jakarta has reduced around 70% of COVID-19 cases spread [20]. A survey reported that 98% of the people in Jakarta have a high perceived severity of COVID-19 and a good compliance towards the mobility restriction regulations [21]. However, the prolonged situation of the pandemic and mobility relaxation might influence people’s compliance towards preventive measures, such as mobility. After the first relaxation period, the trend of mobility in Jakarta rose gradually. When the government of Jakarta implemented the second PSBB in September, reduction in mobility was observed and then increased again after the relaxation was put into effect in October. A study in the United States demonstrated that after the issue of stay-at-home orders in a state of emergency in the early pandemic, the mobility decreased around 35%. However, as the announcement of the reopening and the signs of pandemic fatigue have shown, the mobility rebound rapidly, leading to a positive relationship on COVID-19 infections [22].

Additionally, mobility relaxations and the news regarding the development of COVID-19 vaccines approaching the end of the year might bring a glimpse of hope that might cause risk compensation. Though the long-term effect of the vaccines is still unclear, people could still see this as a silver lining. This whole situation was previously explained as the Peltzman Effect, where individuals respond to safety measures while increasing their risky behavior [23]. The vaccine development for the people might suggest that the pandemic situation is getting somewhat closer to the end, as the herd immunity will be accomplished in the near future. This situation led people to compromise the preventive measures and resulted in risky behavior, such as lower compliance to stay at home and social distancing [23, 24]. Government press release report demonstrated that during public holidays in October, people in Jakarta showed lower compliance in mask-wearing, hand-washing, and social distancing [25].

We support that mobility reduction would not be effective without solid enforcement from the local government. However, we also highlight that populations have the power in terms of their behavioral choice, due to their own perceived risk, to reduce COVID-19 spread by limiting their mobility. Earlier studies reported that due to fears in the H1N1 outbreak in the United States, the public reduced their activity in malls, events, and public transportations [26]. In this study, we assumed that when the mobility declined due to the effect of PSBB, COVID-19 cases should have declined as well. However, the observed COVID-19 cases showed otherwise. COVID-19 cases remained stable while the estimated case started declining. We assumed that instead of transmission in public places, there might be a possibility of household transmission for some time due to the movement limitation. Additionally, during the mobility restriction (PSBB), transmission might not be as high as the relaxation period. Hence the observed graph showed a slightly flat trend.

Moreover, when the mobility limitation was put into effect, the number of cases could not be decreased immediately. Instead, it will turn into a slope first before it finally decreases. Thus, we suggested that other than mobility, other factors are contributing to increased COVID-19 cases.

Limitations of this study are related to the nature of this study, where the analysis using CMR could not solely be the main evidence to explain COVID-19 dynamics. The information on CMR only captures the mobility information from those who have access to smartphones and turn on their location settings. Additionally, recent mobility information on Google CMR represents the data several days prior, making it challenging to utilize for present decision-making purposes. In terms of COVID-19 cases, inadequate testing and delayed reporting numbers could not demonstrate the results in the individual level but the group levels.

Our study utilized only community mobility data to explore the effect of mobility on COVID-19 dynamics, resulting in a partial explanation of the dynamics. Previous studies suggested that compared to other behavioral changes (wearing face masks, hand-washing, social distancing), mobility played a minor role in COVID-19 transmission. Even travel restrictions played a minor role in the spread [27, 28]. Therefore, exploration of other additional drivers of transmission is necessary while analyzing the data. Those contributing factors are including the nature of the virus, public health interventions, and individual behavior [29, 30]. Exploration of indoor and outdoor crowds is also necessary to enhance the analysis, as CMR data does not provide such information.

Furthermore, this study only partially captured the situation at the city level. Jakarta, the biggest city in Indonesia, consists of a very large and dense population. However, the scope of analysis of this study is relatively small, representing only the within-city movement of the population. Moreover, many modes of transportation are available in Jakarta, where inter-city movement should be taken into account to explain COVID-19 dynamics further. Hence, we suggest that a more dynamic approach through model simulation using agent-based modeling will explain the influence of inter-city mobility on COVID-19 dynamics.

## Conclusions

Different types of mobility influence the COVID-19 dynamics. In the events of relaxation and public holidays where mobility is expected to increase, community mobility management and prevention measures will be critical. Particular management should emphasize public places which potentially contribute to increased COVID-19 cases. Decision-makers in public health could utilize the current CMR data analysis to anticipate increased COVID-19 cases in the future, especially during seasonal events such as annual religious holidays or other long holidays.

## Data Availability

This study used the publicly-made available data on COVID-19 and community mobility.
The data on COVID-19 cases data in Jakarta is provided by KawalCOVID-19, an independent website that provides information and collects data on Indonesia's COVID-19 pandemic from the government at the provincial levels.
We retrieved the community mobility data from the Google Community Mobility Report (CMR) Data.

https://kawalcovid19.id/

https://www.google.com/covid19/mobility/

## Acknowledgements

We thank Faraddina Azzahra and Nurul Lathifah for the contribution in the preparation of this manuscript.

## Funding

This work was supported by Directorate of Research, Gadjah Mada University, Indonesia.

## Declaration of interests

We declare no competing interests.

## Notes

### Competing Interest Statement

The authors have declared no competing interest.

### Funding Statement

This work was supported by Directorate of Research, Gadjah Mada University, Indonesia.
The funder of the study had no role in study design, data collection, data analysis, data interpretation, or writing of the report.

### Author Declarations

This study was conducted using a publicly-made available data, therefore IRB approvals is not applicable.

## References

1. John Hopkins. CSSE Coronavirus COVID-19 Global Cases (dashboard) 2020 [Cited 2021 January 15]. Available from: https://gisanddata.maps.arcgis.com/apps/opsdashboard/index.html#/bda7594740fd40299423467b48e9ecf6.

2. Zhang S, Wang Z, Chang R, Wang H, Xu C, Yu X, et al. COVID-19 containment: China provides important lessons for global response. Front. Med. 2020;14(2):215–9. doi: 10.1007/s11684-020-0766-9.

3. Varghese C, Xu W. Quantifying what could have been - The impact of the Australian and New Zealand governments’ response to COVID-19. Infect Dis Health. 2020;25(4):242–4. Epub 2020/06/09. doi: 10.1016/j.idh.2020.05.003.

4. Bajardi P, Poletto C, Ramasco JJ, Tizzoni M, Colizza V, Vespignani A. Human mobility networks, travel restrictions, and the global spread of 2009 H1N1 pandemic. PLoS ONE. 2011;6(1):e16591. doi: 10.1371/journal.pone.0016591.

5. Oliver N, Letouzé E, Sterly H, Delataille S, De Nadai M, Lepri B, et al. Mobile phone data and COVID-19: Missing an opportunity? arXiv preprint arXiv:200312347. [Preprint]. 2020. [Cited 2021 April 12]. Available from: https://arxiv.org/abs/2003.12347v1

6. Badr HS, D. H, Marshall M, Dong E, Squire MM, Gardner LM. Association between mobility patterns and COVID-19 transmission in the USA: a mathematical modelling study. Lancet Infect Dis. 2020;20(11):1247–54. Epub 2020/07/06. doi: 10.1016/s1473-3099(20)30553-3.

7. Asweto CO OP, Alzain MA, Wang W. Effects of increased residential mobility and reduced public spaces mobility in containing covid-19 in Africa. Journal of Global Health Reports. 2020;4(e2020075). doi: doi:10.29392/001c.14152.

8. Sulyok M, Walker M. Community movement and COVID-19: a global study using Google’s Community Mobility Reports. Epidemiol. 2020;148:e284–e. doi: 10.1017/S0950268820002757.

9. Wang HY, Yamamoto N. Using a partial differential equation with Google Mobility data to predict COVID-19 in Arizona. Math Biosci Eng. 2020;17(5):4891–904. Epub 2020/10/31. doi: 10.3934/mbe.2020266.

10. Pérez-Arnal R, Conesa D, Alvarez-Napagao S, Suzumura T, Català M, Alvarez-Lacalle E, et al. Comparative analysis of geolocation information through mobile-devices under different covid-19 mobility restriction patterns in spain. ISPRS Int. J. Geoinf. 2021;10(2):73.

11. Jakarta Response to COVID-19 Outbreak: Jakarta response to covid-19 outbreak: a timeline 2020. 2020 May 29 [Cited 27 April 2021]. Available from: https://corona.jakarta.go.id/en/artikel/linimasa-kebijakan-penanganan-pandemi-covid-19-di-jakarta.

12. KawalCOVID19. Grafik interaktif untuk data per provinsi [internet]. 2020 [Cited 15 January 2021]. Available from: https://kawalcovid19.id/

13. Google LLC. Google COVID-19 Community Mobility Reports. 2020 [Cited 14 January 2021]. Available from: https://www.google.com/covid19/mobility/

14. Praharaj S, Han H. A longitudinal study of the impact of human mobility on the incidence of COVID-19 in India. medRxiv. [Preprint]. 2020 [Cited 15 April 2021]. Available from: https://www.medrxiv.org/content/10.1101/2020.12.21.20248523v1.full2020.12.21.20248523. doi: 10.1101/2020.12.21.20248523.

15. Goolsbee A, Syverson C. Fear, lockdown, and diversion: Comparing drivers of pandemic economic decline 2020. J. Public. Econ. 2021;193:104311. doi: https://doi.org/10.1016/j.jpubeco.2020.104311.

16. Nugraheny DE. Bukan klaster perkantoran, ini penyumbang tingginya kasus covid-19 di DKI versi Satgas. Kompas. 2020 July 29 [Cited 27 April 2021]. [internet]. Available from: https://nasional.kompas.com/read/2020/07/29/11455941/bukan-klaster-perkantoran-ini-penyumbang-tingginya-kasus-covid-19-di-dki.

17. Nishiura H, Oshitani H, Kobayashi T, Saito T, Sunagawa T, Matsui T, et al. Closed environments facilitate secondary transmission of coronavirus disease 2019 (COVID-19). MedRxiv. 2020 [Cited 2021 April 12]. Available from: https://www.medrxiv.org/content/10.1101/2020.02.28.20029272v2

18. Noorimotlagh Z, Jaafarzadeh N, Martínez SS, Mirzaee SA. A systematic review of possible airborne transmission of the COVID-19 virus (SARS-CoV-2) in the indoor air environment. Environ Res. 2021;193:110612-. Epub 2020/12/10. doi: 10.1016/j.envres.2020.110612.

19. Musselwhite C, Avineri E, Susilo Y. Editorial JTH 16 -The coronavirus disease COVID-19 and implications for transport and health. J Transp Health. 2020;16:100853-. Epub 2020/04/04. doi: 10.1016/j.jth.2020.100853.

20. Satyakti Y. Do human restriction mobility policy in Indonesia effectively reduce the spread of COVID-19. MPRA Paper 101911: University Library of Munich, Germany; 2020.

21. Wabah COVID-19: Sikap atas kebijakan dan kondisi ekonomi. Saiful Mujani Research and Consulting, 2020 April 9 [Cited 23 April 2021]. [internet]. Available from: https://saifulmujani.com/wabah-covid-19-sikap-atas-kebijakan-dan-kondisi-ekonomi-warga/

22. Xiong C, Hu S, Yang M, Luo W, Zhang L. Mobile device data reveal the dynamics in a positive relationship between human mobility and COVID-19 infections. PNAS. 2020;117(44):27087–9. doi: 10.1073/pnas.2010836117.

23. Peltzman S. The effects of automobile safety regulation. J. Polit. Econ. 1975;83(4):677–725. doi: 10.1086/260352.

24. Mantzari E, Rubin GJ, Marteau TM. Is risk compensation threatening public health in the covid-19 pandemic? BMJ. 2020;370:m2913. doi: 10.1136/bmj.m2913.

25. Fernando D. People neglect health protocols during holidays: report. The Jakarta Post. 2020 November 5 [Cited 23 April 2021]. [internet]. Available from: https://www.thejakartapost.com/news/2020/11/05/people-neglect-health-protocols-during-holiday-report.html.

26. SteelFisher GK, Blendon RJ, Bekheit MM, Lubell K. The public’s response to the 2009 H1N1 influenza pandemic. NEJM. 2010;362(22):e65. doi: 10.1056/NEJMp1005102.

27. Gatalo O, Tseng K, Hamilton A, Lin G, Klein E. Associations between phone mobility data and COVID-19 cases. Lancet Infect Dis. doi: 10.1016/S1473-3099(20)30725-8.

28. Chinazzi M, Davis JT, Ajelli M, Gioannini C, Litvinova M, Merler S, et al. The effect of travel restrictions on the spread of the 2019 novel coronavirus (COVID-19) outbreak. Science. 2020;368(6489):395–400. Epub 2020/03/08. doi: 10.1126/science.aba9757.

29. Nouvellet PBS; Cori, A; et.al. Reduction in mobility and COVID-19 transmission. Nat. Commun. 2021; 12: 1090.

30. Drake JM, Chew SK, Ma S. Societal learning in epidemics: intervention effectiveness during the 2003 SARS outbreak in Singapore. PLoS One. 2006;1(1):e20. Epub 2006/12/22. doi: 10.1371/journal.pone.0000020.

